# Rapid generation of neutralizing antibody responses in COVID-19 patients

**DOI:** 10.1101/2020.05.03.20084442

**Authors:** Mehul S. Suthar, Matthew G. Zimmerman, Robert C. Kauffman, Grace Mantus, Susanne L. Linderman, Abigail Vanderheiden, Lindsay Nyhoff, Carl Davis, Seyi Adekunle, Maurizio Affer, Melanie Sherman, Stacian Reynolds, Hans P. Verkerke, David N. Alter, Jeannette Guarner, Janetta Bryksin, Michael Horwath, Connie M. Arthur, Natia Saakadze, Geoffrey Hughes Smith, Srilatha Edupuganti, Erin M. Scherer, Kieffer Hellmeister, Andrew Cheng, Juliet A. Morales, Andrew S. Neish, Sean R. Stowell, Filipp Frank, Eric Ortlund, Evan Anderson, Vineet D. Menachery, Nadine Rouphael, Aneesh Mehta, David S. Stephens, Rafi Ahmed, John D. Roback, Jens Wrammert

## Abstract

SARS-CoV-2 is currently causing a devastating pandemic and there is a pressing need to understand the dynamics, specificity, and neutralizing potency of the humoral immune response during acute infection. Herein, we report the dynamics of antibody responses to the receptor-binding domain (RBD) of the spike protein and virus neutralization activity in 44 COVID-19 patients. RBD-specific IgG responses were detectable in all patients 6 days after PCR confirmation. Using a clinical isolate of SARS-CoV-2, neutralizing antibody titers were also detectable in all patients 6 days after PCR confirmation. The magnitude of RBD-specific IgG binding titers correlated strongly with viral neutralization. In a clinical setting, the initial analysis of the dynamics of RBD-specific IgG titers was corroborated in a larger cohort of PCR-confirmed patients (n=231). These findings have important implications for our understanding of protective immunity against SARS-CoV-2, the use of immune plasma as a therapy, and the development of much-needed vaccines.

## INTRODUCTION

COVID-19 is a global pandemic. There is a pressing need to understand the immunological response that mediates protective immunity to SARS-CoV-2. Antibody responses to the spike (S) protein are thought to be to the primary target of neutralizing activity during viral infection, conferring superior protective immunity compared to the membrane (M), envelope (E), and nucleocapsid proteins (Bolles et al., 2011; Buchholz et al., 2004; Deming et al., 2006). The S glycoprotein is a class I viral fusion protein that exists as a metastable prefusion homotrimer consisting of individual polypeptide chains (between 1100–1600 residues in length) responsible for cell attachment and viral fusion (Bosch et al., 2003; Hoffmann et al., 2020; Wang et al., 2020). Each of the S protein protomers is divided into two distinct regions, the S_1_ and S_2_ subunits (Bosch et al., 2003; Tortorici and Veesler, 2019). The S_1_ subunit is a V-shaped polypeptide with four distinct domains, Domains A, B, C, and D, with Domain B functioning as the receptor-binding domain (RBD) for most coronaviruses, including the pathogenic β-coronaviruses such as SARS-CoV-2, SARS and MERS (Fig. 1A; Supplementary Fig. 1A) (Li et al., 2005; Lu et al., 2013; Tortorici and Veesler, 2019; Walls et al., 2020). Recent studies have shown that the SARS-CoV-2 RBD interacts with the ACE2 receptor for cellular attachment (Hoffmann et al., 2020; Walls et al., 2020; Wang et al., 2020). Sequence analysis of the RBD shows extensive homology in this region to SARS (73%). In contrast, MERS and other seasonal coronaviruses show minimal sequence homology to the SARS-CoV-2 RBD (7–18%) (Fig. 1B). Herein, we set out to understand the dynamics, specificity, and neutralizing potency of the humoral immune response against the RBD of the SARS-CoV-2 spike protein during acute infection.

### The magnitude of RBD-specific antibody responses in acutely infected COVID-19 patients

To determine the magnitude of antibody responses, Ig isotype, and IgG subclass usage against the RBD of the SARS-CoV-2 spike protein, we analyzed a cohort of acutely infected COVID-19 patients (n=44) enrolled at two hospitals in the Emory Healthcare System in Atlanta (Emory University Hospital and Emory University Hospital Midtown). These patients were recruited from both the inpatient ward and the ICU (patient details are provided in Table 1). These samples represent a cross-section of days after patient-reported symptom onset (3–30 days) and PCR confirmation (2–19 days). As healthy controls, we used plasma samples collected at baseline in a vaccine study performed in early 2019 (n=12). The RBD protein was cloned and expressed in mammalian cells (Supplementary Fig. 1B) and was validated by ELISA using CR3022, a SARS-specific human monoclonal antibody that cross-reacts with SARS-CoV-2 (ter Meulen et al., 2006) (Supplementary Fig. 1C). Size exclusion chromatography shows that the recombinant RBD protein is homogenous and does not form aggregates (Supplementary Fig. 1D). We found that a majority of COVID-19 patients (36 out of 44) developed RBD-specific class-switched IgG responses (Fig. 1C) (mean titer: 18500, range: <100–142765). These patients also showed IgM and IgA responses of lower magnitude as compared to IgG (IgM mean titer: 3731, range:<100-40197 and IgA mean titer: 973, range: <100–19918). All of the negative controls were below the limit of detection in the endpoint analysis for binding to the RBD antigen (Fig. 1C, red). A representative RBD-specific IgG ELISA assay for a subset of these donors is shown in Figure 1D to illustrate the dynamic range of these measurements. A number of the COVID-19 patient samples that scored either negative or low in the RBD IgG ELISA had higher titers of IgM (Fig. 1E, green). Finally, IgG subclass analysis showed that the COVID-19 patients exclusively made RBD-specific IgG1 and IgG3, with no detectable IgG2 or IgG4. Taken together, these findings illustrate that antibody class-switching to IgG occurs early during acute infection.

**Table 1.** COVID-19 patient cohort.

| Patient ID# | Age | Sex | Days after<br>symptom onset | Days after<br>+ PCR | IgG | IgM | IgA | FRNT50 |
| --- | --- | --- | --- | --- | --- | --- | --- | --- |
| 1 | 65 | M | 8 | 8 | 1692 | 507 | <100 | 761 |
| 2 | 36 | F | 15 | 8 | 86698 | 664 | 313 | 2233 |
| 3 | 41 | M | 8 | 2 | 259 | 599 | 116 | 245 |
| 4 | 44 | F | 9 | 5 | 2560 | 947 | 442 | 194 |
| 5 | 56 | M | 17 | 5 | 10422 | 1574 | 4033 | 3200 |
| 6 | 80 | M | 29 | 7 | 22219 | 1242 | 176 | 2483 |
| 7 | 74 | M | 12 | 2 | <100 | <100 | <100 | <50 |
| 8 | 66 | M | 8 | 3 | 419 | 220 | 85 | 167 |
| 9 | 87 | M | 9 | 3 | <100 | <100 | <100 | 124 |
| 10 | 75 | M | 18 | 6 | 1174 | 369 | 131 | 539 |
| 11 | 61 | M | 19 | 4 | 445 | 282 | <100 | 645 |
| 12 | 80 | M | 5 | 5 | 899 | 470 | 316 | 167 |
| 13 | 66 | M | 10 | 2 | 4988 | 795 | 297 | 1502 |
| 14 | 37 | F | 14 | 3 | 202 | 578 | <100 | 118 |
| 15 | 64 | F | 8 | 3 | <100 | 166 | <100 | 55 |
| 16 | 76 | F | 11 | 2 | 31205 | 2906 | 230 | 1718 |
| 17 | 70 | F | 11 | 2 | 1100 | 246 | <100 | 138 |
| 18 | 63 | F | 12 | 5 | <100 | <100 | <100 | <50 |
| 19 | 70 | M | 5 | 4 | <100 | 136 | <100 | 156 |
| 20 | 66 | M | 22 | 7 | 4269 | 1207 | 324 | 496 |
| 21 | 33 | F | 7 | 3 | 243 | 325 | <100 | 174 |
| 22 | 62 | F | 12 | 7 | 17917 | 4414 | 706 | 603 |
| 23 | 59 | M | 11 | 7 | 21323 | 3865 | 140 | 2799 |
| 24 | 58 | F | 11 | 3 | 317 | 364 | 258 | 175 |
| 25 | 76 | M | 17 | 7 | 28352 | 1493 | 6865 | 2561 |
| 26 | 49 | M | 16 | 3 | 15772 | 491 | 589 | 126 |
| 27 | 54 | M | 30 | 10 | 72949 | 13310 | 700 | 3341 |
| 28 | 73 | M | 11 | 15 | 69902 | 3412 | 19918 | 911 |
| 29 | 64 | F | 9 | 3 | <100 | 119 | 161 | 67 |
| 30 | 37 | F | 13 | 3 | 205 | 419 | <100 | 126 |
| 31 | 25 | F | 3 | 2 | <100 | <100 | <100 | <50 |
| 32 | 39 | F | 8 | 3 | 814 | 362 | 287 | 262 |
| 33 | 60 | M | 10 | 6 | 27557 | 50483 | <100 | 5763 |
| 34 | 60 | F | 15 | 8 | 43072 | 443 | 214 | 337 |
| 35 | 56 | F | 18 | 8 | 71204 | 16298 | 1754 | 1177 |
| 36 | 60 | M | 17 | 12 | 142766 | 40197 | <100 | 5378 |
| 37 | 52 | M | 12 | 5 | <100 | 317 | <100 | 108 |
| 38 | 73 | M | 11 | 4 | 683 | 303 | <100 | <50 |
| 39 | 48 | M | 8 | 6 | 9311 | 1750 | 884 | 1068 |
| 40 | 46 | M | 20 | 13 | 69361 | 706 | 1112 | 408 |
| 41 | 69 | M | 18 | 19 | 33684 | 5517 | 180 | 1882 |
| 42 | 55 | M | 8 | 4 | 377 | 1560 | 178 | 79 |
| 43 | 47 | F | 9 | 3 | 467 | 1042 | <100 | 158 |
| 44 | 63 | F | 10 | 3 | 134 | 186 | <100 | 99 |

**Figure 1.**
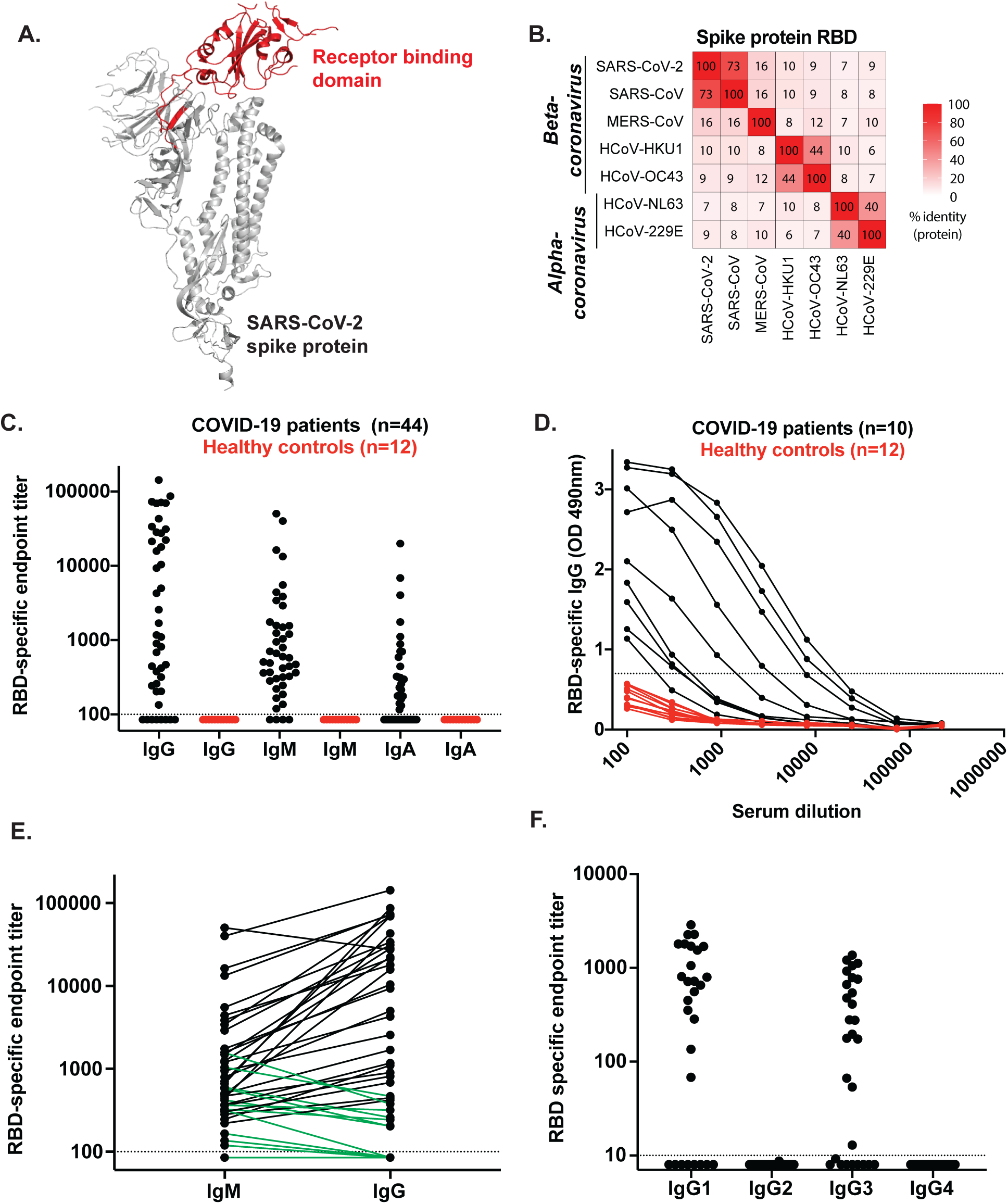
Antibody responses against SARS-CoV-2 RBD in PCR-confirmed acutely infected COVID-19 patients. A) Structure of a SARS-CoV-2 spike protein (single monomer is shown) with the RBD highlighted in red(Wang et al., 2020) B) Sequence homology analysis of SARS-CoV-2 spike protein RBD compared to SARS, MERS, and seasonal alpha- and beta-CoVs. C) ELISA endpoint titers for SARS-CoV-2 RBD specific IgG, IgA and IgM in PCR confirmed acute COVID-19 patients (n=44) and healthy controls collected in early 2019. Endpoint cutoff values were calculated using the mean of the 12 healthy controls at 1/100 dilution, times 3 standard deviations (shown as a dotted line). D) Representative ELISA assays for 10 patients and 12 healthy controls. E) Direct comparison of IgM and IgG for individual donors. A number of the IgG negative or low early samples were IgM positive (shown in green). F) Endpoint titer analysis of IgG subclass distribution. Each experiment was performed at least twice and a representative data set is shown.

### Neutralization potency of antibody responses in COVID-19 patients

We next determined the neutralization capacity of samples from the cohort of acutely infected COVID-19 patients. We have developed a focus reduction neutralization titer (FRNT) assay for SARS-CoV-2. In this assay, COVID-19 patient plasma is incubated with a clinical isolate of SARS-CoV-2 followed by infection of VeroE6 cells (Harcourt et al., 2020). The neutralization potency of the plasma sample is measured by the reduction in virally-infected foci. We screened plasma from COVID-19 patients (n=44) and found that a majority of the samples (40/44) showed neutralization capacity, with titers ranging from 1:5763 to 1:55 (Fig. 2A). A representative example of viral neutralization is shown in Figure 2b where pre-incubation with control plasma yields about 250 foci whereas the COVID-19 patient sample completely inhibited the formation of infected foci (Fig. 2B). Representative neutralization curves for a subset of samples are shown to illustrate the dynamic range of the results obtained (Fig. 2C). A plaque reduction neutralization titer (PRNT) assay is the classic method for determining the neutralization capacity of a plasma sample against coronavirus infection (Rockx et al., 2008). To confirm the efficiency of these two assays, we compared the neutralization titers between a standard PRNT assay and an FRNT assay for a subset of the patient samples (n=9). Overall, we observed a strong positive correlation between these two assays (Fig. 2D), demonstrating the robustness of the FRNT assay Overall, these findings demonstrate that neutralizing antibody responses are generated early during acute COVID-19 infection.

**Figure 2.**
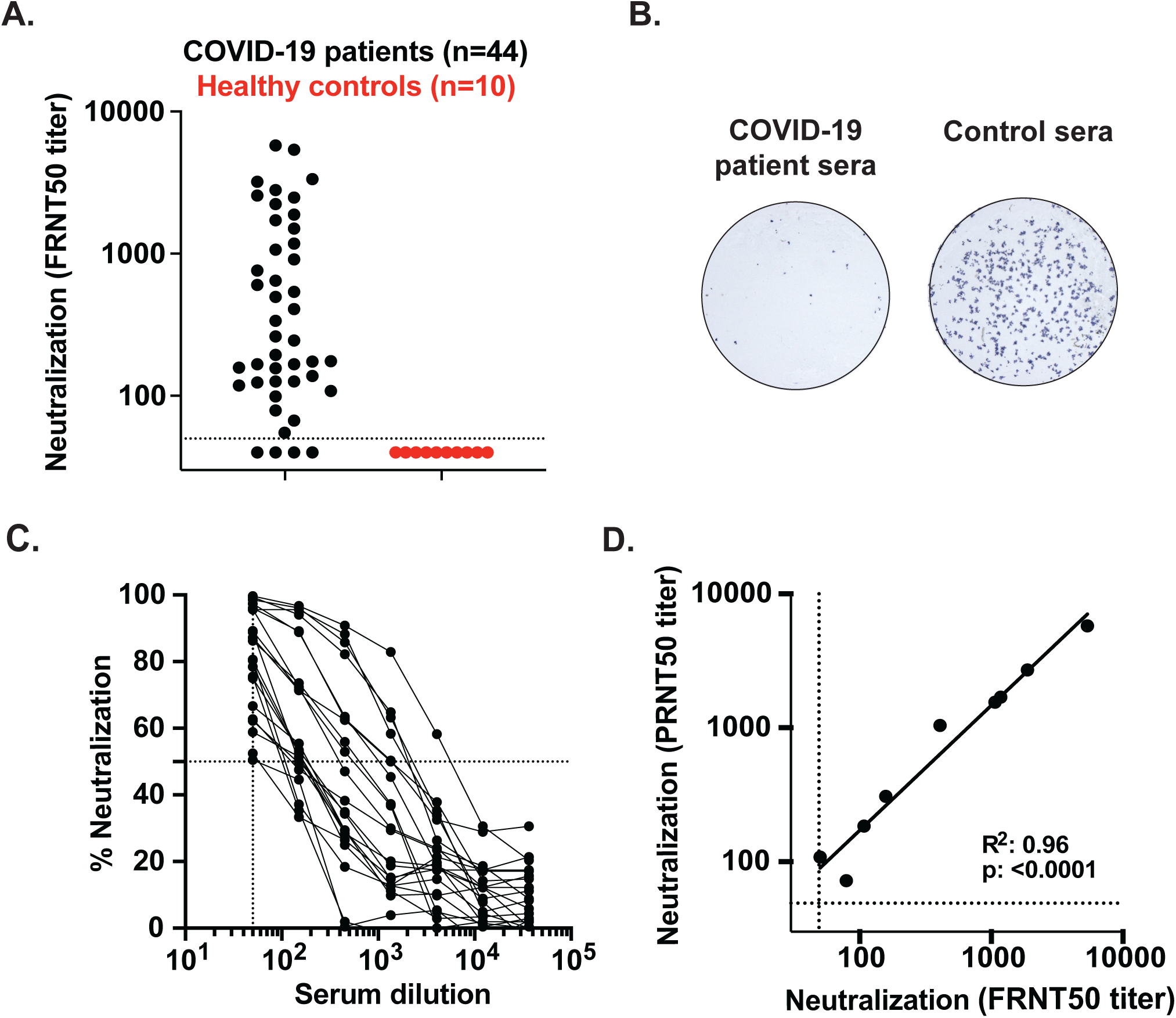
COVID-19 patient plasma neutralizes SARS-CoV-2. A) Neutralization activity of serum samples against SARS-CoV-2. The FRNT_50_ titers of COVID-19 patients (n=44) and healthy controls (n=10) sera were determined by a novel FRNT assay using an immunostain to detect infected foci. Each circle represents one serum sample. The dotted line represents the maximum concentrations of the serum tested (1/50). B) Representative sample showing a reduction in foci from a neutralization assay with sera from an infected COVID-19 patient. C) Representative FRNT_50_ curves (n=22). The dotted line represents 50% neutralization. D) Comparison of PRNT_50_ against FRNT_50_ titers (n=9). Each experiment was performed at least twice and a representative data set is shown.

### Kinetics of the antibody responses during acute SARS-CoV-2 infection

The patient samples were collected across a range of days after symptom onset or PCR confirmation of SARS-CoV-2 infection (Table 1). To understand the relationship between these variables and RBD-specific IgG antibody titers and viral neutralization potency, we performed correlation analyses. In all cases, we observed significant correlations between the number of days elapsed after symptom onset or positive PCR test and the RBD-specific IgG titer or viral neutralization titer (Fig. 3). Several key points regarding the kinetics of antibody responses can be made from this correlation analysis. Antibody responses against the RBD (Fig. 3A), as well as SARS-CoV-2 virus neutralization titers (Fig. 3B), can be detected in a majority of patients around day 8 after symptom onset. When the number of days after PCR confirmation is used to assess the duration of infection, both RBD-binding titers (Fig. 3C) and viral neutralization titers (Fig. 3D) can be detected in many patients already between days 2–6. Beyond 6 days post-PCR confirmation, all patients display both antibody binding and neutralization titers. Taken together, these findings illustrate that both RBD-specific and neutralizing antibody responses occur rapidly after SARS-CoV-2 infection.

**Figure 3.**
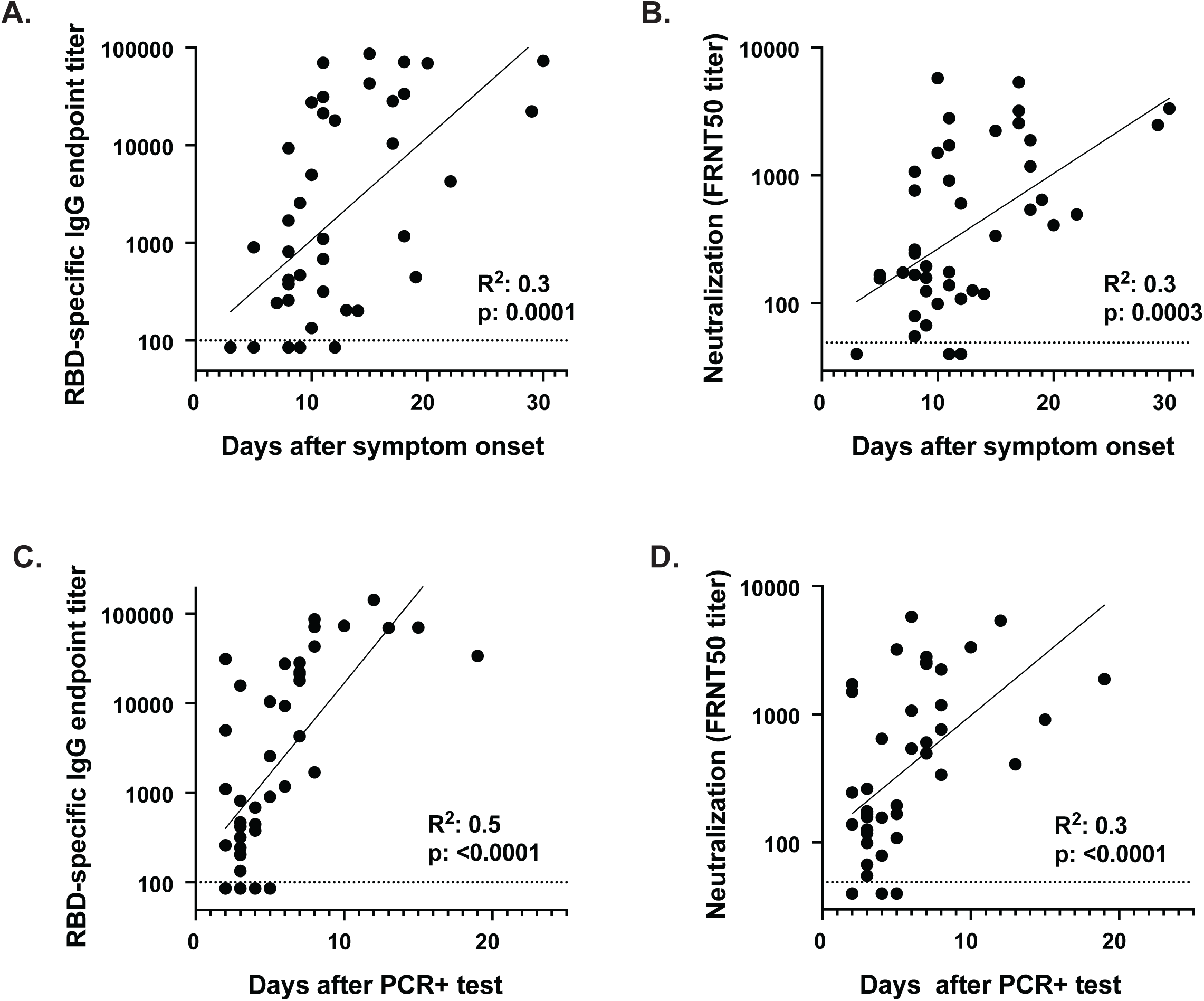
SARS-CoV-2 antibody responses correlate with the progression of acute SARS-CoV-2 infection. Comparison of RBD-specific IgG titers and neutralization titers with (A-B) days after symptom onset or (C-D) days after PCR positive confirmation for each patient. Correlation analysis was performed by log transformation of the endpoint ELISA titers followed by linear regression analysis.

### RBD-specific antibody titers as a surrogate of neutralization potency in acutely infected COVID-19 patients

We observed a wide range of RBD-specific and neutralizing antibody responses across the cohort of acutely infected COVID-19 patients. We found that the magnitude of RBD-specific IgG titers positively correlated with neutralization titers (r^2^= 0.7; *p*<0.0001; Fig. 4A). Overall, we observed viral neutralization activity in 40 out of 44 samples from acutely infected COVID-19 patients.

**Figure 4.**
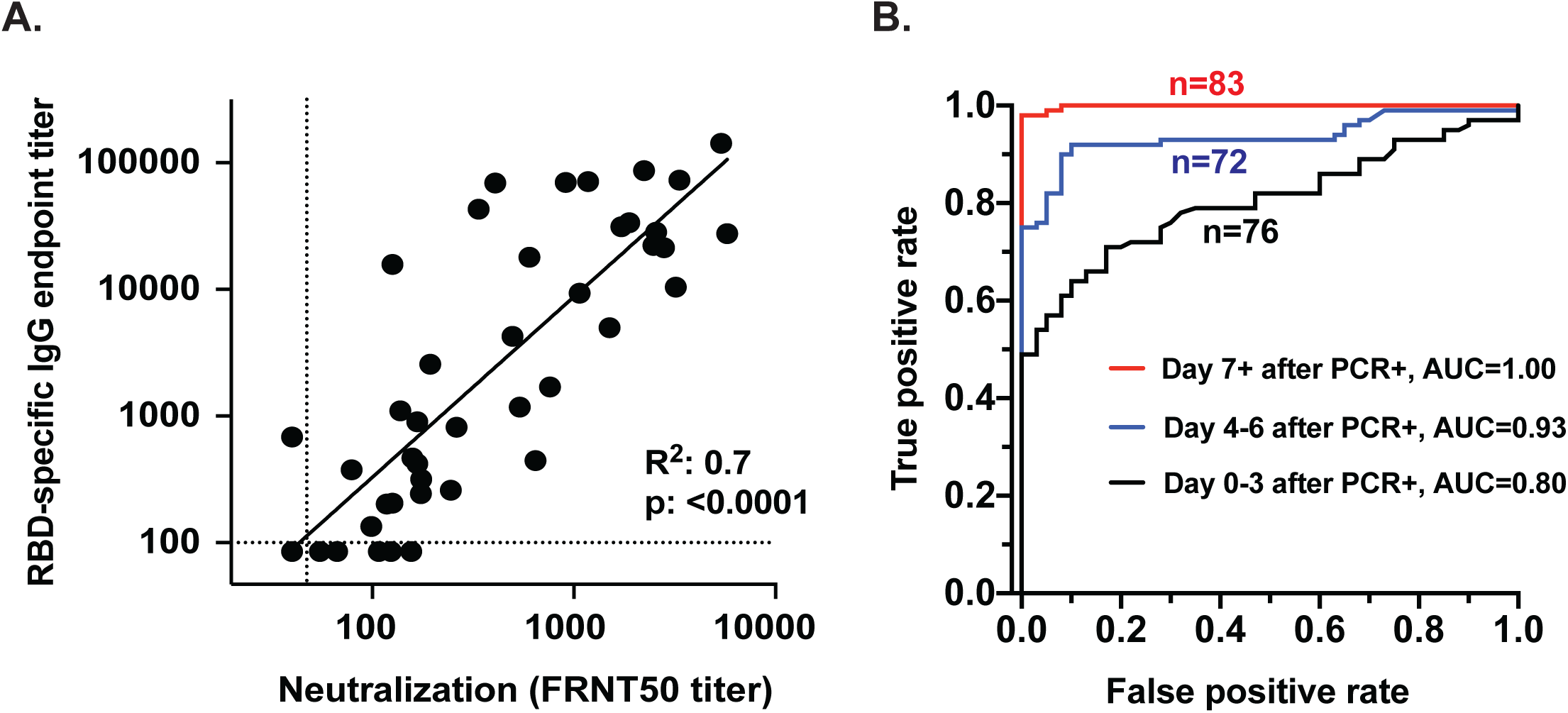
RBD-specific antibody titers as a surrogate of neutralization potency in acutely infected COVID-19 patients. A) Comparison of RBD-specific IgG endpoint titers with SARS-CoV-2-specific FRNT_50_ titers. Correlation analysis was performed by log transformation of the endpoint ELISA or FRNT_50_ titers followed by linear regression analysis. B) The RBD-specific ELISA was validated for high-throughput clinical testing in Emory Medical Laboratories. Sera (n=231) were collected from COVID-19 patients within the first 22 days after PCR-confirmation (Supplementary Table 1). Sera (n=40) collected in 2019 were used as negative controls. ROC curves are shown comparing the true positive and false negative rates of the ELISA using different OD cutoffs and sera collected at different times post-infection. Whereas the RBD ELISA produced an area under the curve (AUC) of 0.80 when samples were collected close to the time of infection (within 3 days of positive PCR; n=76), longer sampling times resulted in better performance. Assay performance was nearly perfectly discriminatory (AUC = 1.00) when samples were collected at least 7 days after the positive PCR (n=83).

We next validated the RBD-specific IgG ELISA for high-throughput testing at the Emory Medical Laboratories. For these analyses, we collected serum from 231 PCR-confirmed COVID-19 patient samples within the first 22 days after PCR confirmation (Supplementary Table 1). In addition, 40 samples collected in 2019 were used as negative controls. These samples were grouped from 0–3 days, 4–6 days, and 7 or more days after PCR confirmation and analyzed using a high-throughput clinical RBD ELISA. The cumulative results of these efforts are shown as receiver operating characteristic (ROC) curves (Fig. 4B). This assay is almost perfectly discriminatory on day 7 or later after PCR confirmation, with an area under the curve (AUC) of 1.00 (n=83). When utilized earlier in the disease course, the performance of this diagnostic assay is reduced. When the RBD specific IgG ELISA were analyzed for the samples collected closer to the time of infection, the AUC for the day 4–6 group (n=76) and the day 0–3 group (n=72) fell to 0.93 and 0.80, respectively. Using an OD cut-off of 0.175 resulted in calculated sensitivity and specificity values of 97.5% and 98%, respectively. Taken together, these findings demonstrate that RBD-specific IgG titers could be used as a surrogate of neutralization activity against SARS-CoV-2 infection and that the RBD assay is highly specific and sensitive. Further, this demonstrates the necessity of appropriate timing of sample collection when using serologic diagnostic tests of acutely infected COVID-19 patients (Lee et al., 2020; Okba et al., 2020).

## DISCUSSION

In this study, we show that RBD-specific IgG antibody responses are rapidly induced during severe and moderate acute COVID-19 infection, with most patients showing RBD-specific antibody responses by 6 days post-PCR confirmation. Consistently, we found that class-switching also occurs early during infection and is dominated by RBD-specific IgG1 and IgG3 responses. We also detected both RBD-specific IgM and IgA responses at relatively lower levels as compared to IgG. These responses result in neutralizing antibody responses that directly correlated with RBD-specific IgG antibody titers. These findings strongly indicate that a robust humoral immune response occurs early during severe or moderate COVID-19 infections.

The validation of the sensitive and selective RBD based clinical assay at EML and the correlation with viral neutralization is promising for both diagnostic purposes and ongoing seroprevalence studies of healthcare workers and the general population. These serology tests could be used for making informed decisions for convalescent plasma therapy that are currently undergoing clinical testing as a possible therapeutic or even prophylactic option (Bloch et al., 2020; Shen et al., 2020). Further, the kinetic findings presented herein are essential for ongoing efforts aimed at applying antibody testing for clinical diagnostic purposes, highlighting the importance of appropriate timing of these tests relative to PCR testing and/or symptom onset after infection. A comprehensive understanding of the dynamics of antibody responses after infection will also be key for understanding disease pathogenesis, risk assessment in vulnerable populations, evaluation of novel therapeutics, and development of vaccines.

The appearance of high titer neutralizing antibody responses early after the infection is promising and may offer some degree of protection from re-infection. Future studies will need to define the neutralizing titer which constitutes a robust correlate of protective immunity and determine the durability of these responses over time (Liu et al., 2006). This information will be essential for ongoing vaccine development efforts (Amanat and Krammer, 2020).

## Data Availability

The data that support the findings of this study are available from the authors on reasonable request, see author contributions for specific data sets.

## Acknowledgments

We would like to thank Laurel Bristow, Ariel Kay, Youssef Saklawi, Ghina Alaaeddine, Nina McNair, Ellie Butler, Brandi Johnson, Christopher Huerta, Jennifer Kleinhenz, Vinit Karmali, Yong Xu, Dongli Wang, Michele McCullough for sample processing at the Hope Clinic. We also acknowledge the dedicated efforts of Hassan Bilal, DeAndre Brown, Davette Campbell, Lisa Cole, Ginger Crews, Shanessa Fakour, Natalie Hicks, Mark Meyers, and Katherine Normile for sample collection, processing, and organization. We thank Gabrielle Holenstein, Corin Jones, Alethea Luo-Gardner, Hoa Nguyen, Keyanna Seville, and Corazon Tomblin for the exceptional technical performance of the ELISA in EML. We also acknowledge thorough and rapid chart reviews by Kari Broder. Finally, we thank all the participating patients, and the hospital staff caring for them.

## Author contributions

M.S, S.R, H.P.V, D.N.A, J.G, G.M, S.L, A.V contributed to the acquisition, analysis, and interpretation of the data, J. B and M.C.H contributed to the acquisition and interpretation of the data, C.M.A and N.S. contributed to the acquisition of the data, G.H.S, M.G.Z, R.C.K contributed to the acquisition, analysis, and interpretation of the data, and helped draft the work, A.M, A.S.N and S.R.S contributed to the acquisition, analysis, and interpretation of the data, as well as the conception and design of the work, D.S.S, L.N, S.A, M.A, and V.D.M contributed to the analysis, and interpretation of the data, S.E served as the principal investigator of the clinical protocol for acquisition of patient samples and contributed to the interpretation of data, E.S, K.H, A.C, J.A.M, N.R, and E.A contributed to the acquisition and interpretation of the data, J.D.R, R.A, J.W, and M.S.S contributed to the acquisition, analysis, and interpretation of the data, as well as the conception and design of the work, and writing the manuscript.

## STAR Methods

### CONTACT FOR REAGENT AND RESOURCE SHARING

Further information and requests for resources and reagents should be directed to and will be fulfilled by the corresponding authors Mehul Suthar and Jens Wrammert.

### EXPERIMENTAL MODEL AND SUBJECT DETAILS

**Ethics statement**. The serum and plasma samples used for this study were collected at Emory University Hospital and Emory University Hospital Midtown in Atlanta. All patients were diagnosed with acute SARS-CoV-2 infection by PCR, and samples were collected at a range of times post-PCR-confirmation. All collection, processing, and archiving of human specimens was performed under approval from the University Institutional Review Board (IRB #00000510 and #00022371). For IRB #00000510, informed consent was obtained prior to patient participation. For #00022371, an IRB waiver was obtained allowing the use of discarded samples in the clinical laboratory at the Emory Hospital.

**Virus and cells**. SARS-CoV-2 (2019-nCoV/USA_WA1/2020) was isolated from the first reported case in the US (Harcourt et al., 2020). A plaque purified passage 4 stock was kindly provided by Natalie Thornburg (CDC, Atlanta, GA). Viral titers were determined by plaque assay on Vero cells (ATCC). Vero cells were cultured in complete DMEM medium consisting of 1x DMEM (Corning Cellgro), 10% FBS, 25mM HEPES Buffer (Corning Cellgro), 2mM L-glutamine, 1mM sodium pyruvate, 1x Non-essential Amino Acids, and 1x antibiotics.

**Cloning, expression, and purification of SARS-CoV-2 RDB**. A recombinant 347 form of the spike glycoprotein receptor-binding domain (RBD) from SARS-CoV-2, Wuhan-Hu-1 (GenPept: QHD43416) was cloned for mammalian expression in human embryonic kidney expi293F cells. The receptor-binding domain consisting of amino acids 319 (arginine) to 541 (phenylalanine) of the SARS-Cov-2 S gene was amplified by PCR using a mammalian codon-optimized sequence as the DNA template (Genscript MC_0101081). PCR amplification appended the first 12 amino acids of the native S gene signal peptide sequence to the N-terminal end of the protein and, at the C-terminal end a 6X polyhistidine tag preceded by a short linker sequence (GGGGS). Forward and Reverse primer sequences consisted were: 5’-AGAGAATTCACCATGTTCGTCTTCCTGGTCCTGCTGCCTCTGGTCTCCAGGGTGCAGCCACCGAGTCTATC-3’ and 5’-CTCTAAGCTTCTATCATTAGTGGTGGTGGTGGTGGTGGCTTCCGCCTCCGCCGAAGTTCACGCACTTGTTCTTCAC-3’. 25 uL PCR reaction conditions were: 1X Phusion HF Buffer, 0.2 mM dNTP, 0.63 units Phusion DNA polymerase, and 500 nM of each primer. PCR cycling conditions were: initial denaturation at 98°C, 1 minute; then 25 cycles of: 98°C, 20 seconds, 65°C 30 seconds, 72°C 30 seconds; followed a final extension at 72°C for 5 minutes. Following amplification, purified PCR products (QIAquick PCR Purification, Qiagen) were digested with EcoRI-HF (NEB) and HindIII (NEB) and cloned into the EcoRI-HindIII cloning site of a mammalian expression vector containing a CMV promoter (Genbank Reference ID FJ475055). Plasmid DNA was prepared using the Qiagen PlasmidPlus Midi purification system and constructs were sequence verified. Recombinant protein expression was performed in Expi293F cells according to the manufacturer’s instructions (Thermo Fisher Scientific). Briefly, expression plasmid DNA was complexed with the expifectamine lipid-based transfection reagent. Complexes were added to the cell suspensions shaking at 125 RPM and incubated overnight at 37°C in an 8% CO_2_ humidified incubator. After 20 hours, protein expression supplements and antibiotics were added. Cultures were then incubated for an additional three days to allow for expression into the supernatant. Cell culture supernatants were harvested by centrifugation at 16,000xg for 10 minutes. Supernatants were sterile filtered through a 0.2 um filter and stored at 4°C for <7 days before purification. Analytical SDS-PAGE was performed on supernatants and the protein concentration in solution was determined by densitometry relative to the purified protein. Recombinant RBD protein levels were between 100 mg and 150 mg per liter. Purification was performed according to manufacturer’s instructions using 5 mL HisTALON Superflow Cartridges (Clontech Laboratories). Briefly, an additional 11.7 g/L of sodium chloride and 0.71 g/L of cobalt(II) chloride hexahydrate were added to culture supernatants, which were adjusted to pH 7.5. The supernatant was then loaded on to the column equilibrated with 10 column volumes of 50 mM phosphate 300 mM sodium chloride buffer pH 7.5 (equilibration buffer). The column was washed with 8 column volumes of equilibration buffer supplemented with 10 mM imidazole. Protein was eluted with 6 column volumes of equilibration buffer supplemented with 150 mM imidazole. The eluted protein was dialyzed overnight against 80 volumes of phosphate-buffered saline pH 7.2. The protein was filter-sterilized (0.2 μm) and normalized to 1 mg/mL by UV spectrophotometry using an absorption coefficient of 1.3 AU at 280 nm=1 mg/mL. Proteins were aliquoted and stored at −80°C prior to use. SDS-PAGE analysis of purified recombinant protein stained with coomassie blue demonstrated that samples were >90% pure (Fig. 1). The RBD resolves at an apparent molecular weight of 30 kDa (Fig. 1D) which is slightly larger than the theoretical molecular weight of 26.5 kDa, presumably caused by glycosylation.

**Preparation of CR3022 monoclonal antibody and biotinylation**. The SARS-CoV S glycoprotein specific antibody CR3022 was generated recombinantly using previously reported heavy and light variable domain sequences deposited in GenBank under accession numbers DQ168569 and DQ168570(ter Meulen et al., 2006). Antibody variable domain gene sequences were synthesized by IDT and cloned into human IgG1 and human kappa expression vectors as previously described (Smith 2009). Antibodies were produced in Expi293F cells according to the manufacturer’s recommendations by co-transfecting heavy and light chain plasmids at a ratio of 1:1.5. Antibodies were purified using rProtein A Sepharose Fast Flow antibody purification resin (GE Healthcare) and buffer exchanged into PBS before use. Biotinylated versions of CR3022 used in viral neutralization assays were produced by combining the antibody with a 20 molar excess of EZ-Link NHS-PEG4-Biotin (Thermo Fisher Scientific) for 1 hour at room temperatures. Reactions were stopped by adding Tris pH 8 to a final concentration of 10 mM. The biotinylated antibody was then buffer exchanged >1000X into PBS using a 10 kDa protein spin-concentrator (Amicon).

## Sequence analysis and alignment

The SARS-CoV-2 spike protein structure(Wrapp et al., 2020) was visualized in Pymol (Schrödinger, LLC). To assess the homology of coronavirus spike proteins, a global protein alignment was performed in Geneious (Geneious, Inc.) with translations of genome sequences accessed through NCBI Nucleotide. Sequences used were GenBank MN908947.3 (SARS-CoV-2), RefSeq NC_004718.3 (SARS-CoV), RefSeq NC_019843.3 (MERS-CoV), NC_006577.2 (HCoV-HKU1), RefSeq NC_006213.1 (HCoV-OC43), RefSeq NC_005831.2 (HCoV-NL63), and RefSeq NC_005831.2 (HCoV-229E). Homology at the RBD was determined by sequence identity between SARS-CoV-2 RBD residues T302 to L560 (Walls et al., 2020; Wang et al., 2020).

**ELISA assays**. Recombinant SARS-CoV-2 RDB was coated on Nunc MaxiSorp plates at a concentration of 1 μg/mL in 100 uL phosphate-buffered saline (PBS) at 4°C overnight. Plates were blocked for two hours at room temperature in PBS/0.05%Tween/1% BSA (ELISA buffer). Serum or plasma samples were heated to 56°C for 30 min, aliquoted, and stored at −20°C before use. Samples were serially diluted 1:3 in dilution buffer (PBS-1% BSA-0.05% Tween-20) starting at a dilution of 1:100. 100 μL of each dilution was added and incubated for 90 minutes at room temperature. 100 uL of horseradish peroxidase-conjugated isotype and subclass specific secondary antibodies, diluted 1 to 2,000 in ELISA buffer, were added and incubated for 60 minutes at room temperature. Development was performed using 0.4 mg/mL o-phenylenediamine substrate (Sigma) in 0.05 M phosphate-citrate buffer pH 5.0, supplemented with 0.012% hydrogen peroxide before use. Reactions were stopped with 1 M HCl and absorbance was measured at 490 nm. Between each step, samples were washed four times with 300 uL of PBS-0.05% Tween. Prior to development, plates were additionally washed once with 300 uL of PBS. Secondary antibodies used for development were as follows: anti-hu-IgM-HRP, anti-hu-IgG-HRP, and anti-hu-IgA-HRP (Jackson Immuno Research, and Mouse anti-hu-IgG1 Fc-HRP, Mouse anti-hu-IgG2 Fc-HRP, Mouse anti-hu-IgG3 Fc-HRP, or Mouse anti-hu-IgG4 Fc-HRP (Southern Biotech).

**Clinical RBD ELISA assay**. This assay was performed essentially as described above, with the following modifications to increase throughput: all serum samples were diluted 1:200, and the incubation times were reduced to 30 minutes after the addition of serum samples and the secondary antibody conjugate.

**Focus Reduction Neutralization Assays**. Serially diluted patient plasma and COVID-19 (100-200 FFU) were combined in DMEM + 1% FBS (Corning Cellgro), and incubated at 37°C for 1 hour. The antibody-virus mixture was aliquoted on a monolayer of VeroE6 cells, gently rocked to distribute the mixture evenly, and incubated at 37°C for 1 hour. After 1 hour, the antibody-virus inoculum was removed and prewarmed DMEM supplemented with 1% FBS (Optima, Atlanta Biologics), HEPES buffer (Corning Cellgro), 2mM L-glutamine (Corning Cellgro), 1mM sodium pyruvate (Corning Cellgro), 1x Non-essential Amino Acids (Corning Cellgro), 1x antibiotics (penicillin, streptomycin, amphotericin B; Corning Cellgro) was mixed with methylcellulose (DMEM [Corning Cellgro], 1% antibiotic, 2% FBS, 2% methylcellulose [Sigma Aldrich]) at a 1:1 ratio and overlaid on the infected VeroE6 cell layer. Plates were incubated at 37°C for 24 hours. After 24 hours, plates were gently washed three times with 1x PBS (Corning Cellgro) and fixed with 200 μl of 2% paraformaldehyde (Electron Microscopy Sciences) for 30 minutes. Following fixation, plates were washed twice with 1x PBS and 100 μl of permeabilization buffer (0.1% BSA-Saponin in PBS) (Sigma Aldrich), was added to the fixated Vero cell monolayer for 20 minutes. Cells were incubated with an anti-SARS-CoV spike protein primary antibody conjugated to biotin (CR3022-biotin) for 1–2 hours at room temperature, then with avidin-HRP conjugated secondary antibody for 1 hour at room temperature. Foci were visualized using True Blue HRP substrate and imaged on an ELISPOT reader (CTL). Each plate contained three positive neutralization control wells, three negative control wells containing healthy control serum mixed with COVID-19, and three mock-infected wells.

## QUANTIFICATION AND STATISTICAL ANALYSIS

**Statistical analysis**. FRNT_50_ curves were generated by non-linear regression analysis using the 4PL sigmoidal dose curve equation on Prism 8 (Graphpad Software). Maximum neutralization (100%) was considered the number of foci counted in the wells infected with a virus mixed with COVID-19 naïve healthy patient serum. Neutralization titers were calculated as 100% × [1-(average number of foci in wells incubated with COVID-19 patient serum) ÷ (average number of foci in wells incubated with control serum)]. For the clinical data Receiver Operating Characteristic (ROC) curves were generated separately for each of three cohorts of clinical validation samples with progressively increasing PCR-to-serum collection intervals (0–3 days, 4–6 days, 7+ days). Optical densities (OD) for each sample were entered into Microsoft Excel for Office 365 v16, and a custom software package was used to iteratively compute the false positive rate (1 - specificity) and true positive rate (sensitivity) at every OD cutoff level for each cohort. The false-positive rates (x) and true positive rates (y) were then rendered as scatter plots to generate the ROC curves. Correlations analyses were done by log transforming RBD binding titers or neutralization titers, followed by linear regression analysis. The R^2^ and p-value are reported in each figure.

## Supplemental Information

**Supplementary Figure 1.**
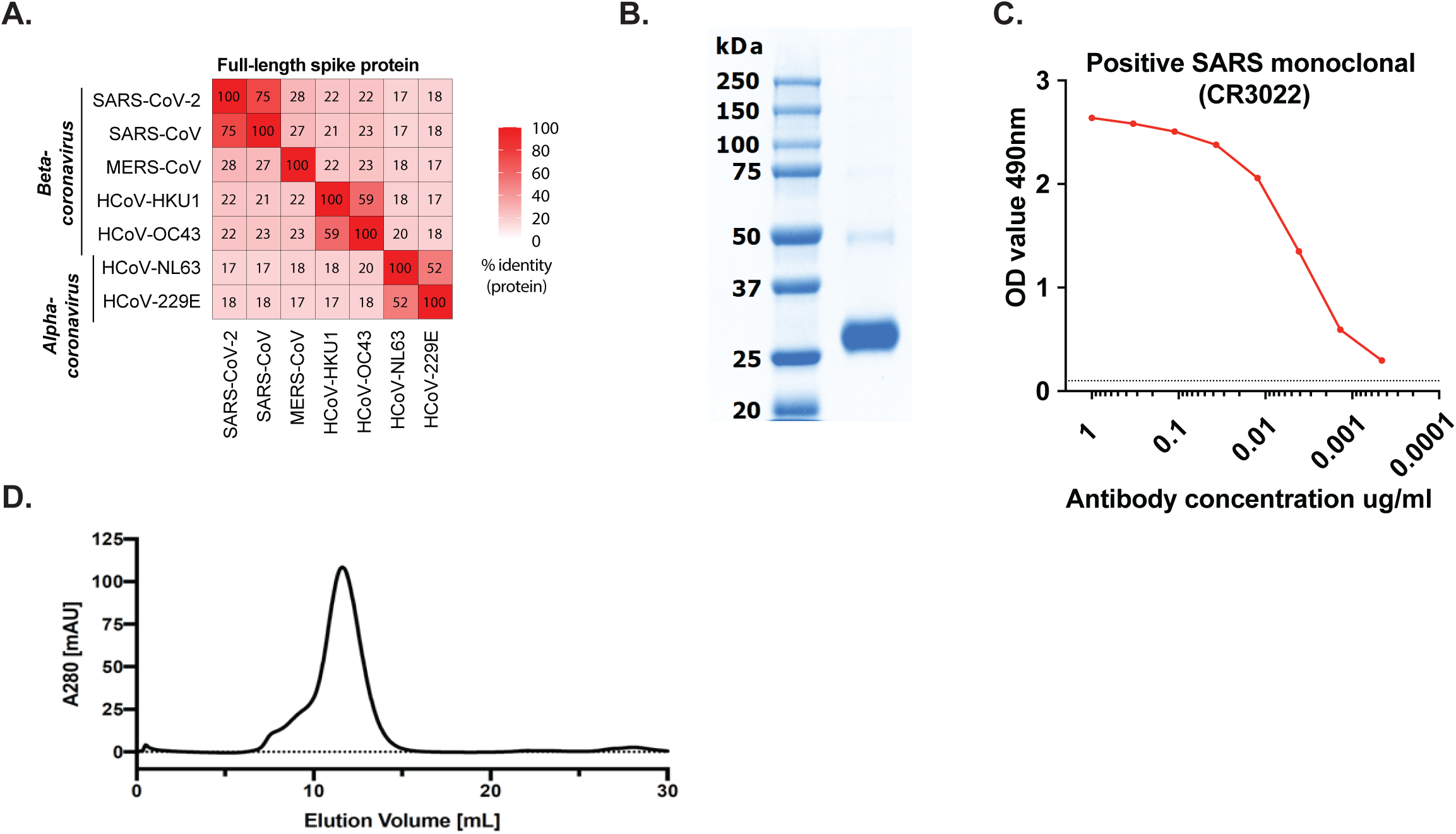
Expression and purification of SARS-CoV-2 RBD. A) Sequence homology analysis of the full-length SARS-CoV-2 spike compared to SARS, MERS, and seasonal alpha- and beta-CoVs. B) SDS-PAGE gel of purified SARS-CoV-2 RBD, cloned and expressed in Expi293F cells by transient transfection. C) ELISA validation of the RBD protein using a monoclonal antibody (CR3022) (ter Meulen et al., 2006) directed against the spike protein RBD. D) Size exclusion chromatography of the recombinant RBD protein. The figure shows the elution profile (UV absorption 280 nm) of 1 mg RBD protein analyzed in PBS buffer on a Superdex 75 (10/300) size exclusion column.

**Supplementary Table 1.** Emory Medical Laboratories patient cohort (time after PCR-confirmation)

| Day post PCR confirmation | Number of cases (n) | Grouping (Days) | Total number |
| --- | --- | --- | --- |
| 0 | 12 | 0-3 | **n=76** |
| 1 | 14 | 0-3 |  |
| 2 | 20 | 0-3 |  |
| 3 | 30 | 0-3 |  |
| 4 | 21 | 4-6 | **n=72** |
| 5 | 29 | 4-6 |  |
| 6 | 22 | 4-6 |  |
| 7 | 15 | 7+ | **n=83** |
| 8 | 14 | 7+ |  |
| 9 | 7 | 7+ |  |
| 10 | 4 | 7+ |  |
| 11 | 4 | 7+ |  |
| 12 | 4 | 7+ |  |
| 13 | 3 | 7+ |  |
| 14 | 2 | 7+ |  |
| 15 | 4 | 7+ |  |
| 16 | 4 | 7+ |  |
| 17 | 4 | 7+ |  |
| 18 | 6 | 7+ |  |
| 19 | 3 | 7+ |  |
| 20 | 5 | 7+ |  |
| 21 | 2 | 7+ |  |
| 22 | 2 | 7+ |  |

